# The primary care annual dementia review: a qualitative study of the views and experiences of service users and providers

**DOI:** 10.1101/2022.04.26.22274255

**Authors:** Alison Wheatley, Greta Brunskill, Johanne Dow, Claire Bamford, Marie Poole, Louise Robinson, the PriDem study team

## Abstract

**Background:** In England and Wales, the Quality and Outcomes Framework (QOF) financially rewards GP practices for long-term conditions management, including completion of annual dementia reviews. There is limited evidence about how this works in practice and whether it meets patients’ and carers’ needs.

**Methods:** Data from five qualitative datasets were integrated and analysed thematically. Data comprised interviews, focus groups, and observations with 209 participants, including commissioners, managers and frontline staff in dementia services; people with dementia; carers; and policy experts.

**Findings:** Four main themes were developed: (i) perceived benefits of annual review; (ii) variability and (in)visibility of annual review; (iii) logistics; and (iv) external influences and constraints.

Variability in both the completion and quality of QOF annual dementia reviews was attributed by some to limited nuance in the current QOF dementia indicator. Many patients and carers were unaware that an annual dementia review had occurred. Participants suggested that many GPs lack the required competencies and/or capacity for successful dementia reviews.

**Conclusions:** Work is urgently needed to improve the quality of annual dementia reviews. Potential strategies include changing the financial reimbursement to reflect both quality and quantity, so the review is tailored to the needs of the individual and their family; the creation of standardised templates; collaborative working within primary care and across sectors; and integrating dementia reviews into other long-term conditions.

**Key points:**

- QOF annual dementia reviews are a key opportunity for providing support for people with dementia in England and Wales
- Current provision of annual reviews varies both in completion rates and quality
- Strategies for improvement include improving quality indicators, implementing standardised templates, and improving primary care capacity and capability to carry out reviews

## Introduction

Dementia is a global public health challenge, with policy currently focused around prevention, timely diagnosis, and improving post-diagnostic support [1, 2]; the latter is the focus of 2022 World Alzheimer Report. Previous World Alzheimer Reports [3, 4] have strongly urged a task-shifted and task-shared approach to post-diagnostic support, with greater involvement of primary care in line with other long-term condition (LTC) management, to cope with increasing demand from our ageing populations. Such primary care involvement has already begun in England and Wales [5, 6], where prevention and management of LTCs, including dementia, are monitored and incentivised via the Quality and Outcomes Framework (QOF)[7]. QOF was introduced in 2004 and links GP financial reimbursement to organisational quality indicators. Reviews of the overall impact of QOF have found positive effects such as reduced mortality rates, better data recording, improved sociodemographic inequalities, [8] and reduction in emergency admissions [9]. Potential problems include poorer quality of care in activities that are not incentivised and poor patient experiences due to ‘tick box exercises’ [8].

QOF dementia indicators have changed over time and currently comprise: (i) the establishment and maintenance of a register of patients diagnosed with dementia; and (ii) a face-to-face review of their care plan at least annually (hereafter, ‘annual dementia review’). Recent research in England and Wales revealed considerable inequalities in post-diagnostic dementia care and support [10-12] and a lack of shared understanding amongst professionals around the duration and nature of post-diagnostic support [13]; the annual dementia review is therefore a major opportunity for monitoring care and providing ongoing support beyond the immediate post-diagnostic period. Although there is some guidance about the content of these reviews (e.g. physical, mental health and social reviews; recording the patients’ future wishes; and identification of carer(s)), financial reimbursement is based on whether or not annual dementia reviews are completed and varies according to the proportion of people living with dementia who receive a review. A GP practice must complete at least 35% of its reviews to receive the minimum payment and 70% for the maximum payment [7].

However, in contrast with other conditions, little attention has been given to the dementia QOF and the implementation of annual dementia reviews in practice. In the 2018/19 financial year, NHS data showed that 95% of GP practices participated in the QOF; of these 97% received the maximum number of points available for dementia (i.e. fully addressing indicators) [14]. This is the most recent information available, as during COVID-19 the QOF was temporarily paused, with many indicators calculated on past performance [15, 16].

Previous studies reviewing patient notes indicate lower proportions of people living with dementia with a documented annual dementia review than published NHS figures suggest [17, 18]. Other research observes that there are potential benefits of annual dementia reviews, including reduced risk of admission to hospital admission [19] or care homes [20]; however, the consistent evidence from families affected by dementia that they experience a lack of post-diagnostic support and care [10-12] suggests that annual dementia reviews may not be adequately addressing their needs.

This paper explores the perspectives of people living with dementia and carers (service users) and professionals (service providers, commissioners, and policy makers) on the annual dementia review, including experiences of review, perceived benefits, and factors influencing the delivery of good quality dementia reviews in primary care.

## Methods

This paper brings together data on annual dementia reviews from multiple qualitative datasets connected to the ‘Primary care led post-diagnostic dementia care: developing evidence-based, person-centred sustainable models for future care’ (PriDem) study [21], which aims to investigate post-diagnostic dementia support in England and Wales. PriDem has multiple workstreams (WS) and a linked postgraduate Annual Dementia Review (ADR) project [22], each of which is described in Table 1. The methods for WS2.1 [13], WS2.2 [23], WS3.1 [24] and WS3.2 [25] are reported in more detail elsewhere. Data collection in PriDem was primarily focused on post-diagnostic care overall but included questions relating to management of dementia in primary care and dementia annual reviews.

**Table 1:** Description of qualitative datasets

|  | WS2.1 | WS2.2 | WS3.1 | WS3.2 | ADR |
| --- | --- | --- | --- | --- | --- |
| Aim | To map the national picture of post-diagnostic support | To explore current examples of good practice in depth | To gain feedback on the developing PriDem intervention | To explore professionals' experiences of post-diagnostic support during COVID-19 | To explore views on the annual dementia review in primary care |
| Participants | Professionals responsible for commissioning and developing dementia services (hereafter 'commissioners'); service managers | Service managers; frontline staff; people living with dementia; and carers | Commissioners; service managers; frontline staff; experts; people living with dementia; and carers | Commissioners; service managers; and frontline staff | Professionals with key roles in shaping dementia policy and national and local dementia champions (hereafter 'experts') |
| Sampling | Dementia services identified through internet searches, documentation (e.g. good practice guidance, award winners), and a survey[26] of commissioners in England | Six services identified during WS2.1 selected for further in-depth case study | Participants from WS2.1 and WS2.2 | Participants from WS2.1 and WS2.2, or colleagues where original participants were unavailable | Participants identified through committee lists, publications and websites, and via project collaborators. |
| Data collection methods | Telephone or face-to-face semi-structured interviews and focus groups | Face-to-face semi-structured interviews, focus groups and observation | Online task groups | Telephone or online semi-structured interviews | Telephone or face-to-face semi-structured interviews |
| Data collection instruments | Topic guide explored: views on task-shifting dementia care to primary care; benefits, facilitators and barriers; and support primary care would need. | Service user topic guide explored: views and experiences of annual dementia review including whether they had one; what they liked; what could be improved; timing and frequency; and whether the review was tailored. Professional topic guide explored: links with primary care including MDTs and shared patient records. | Topics covered included feasibility of implementing PriDem intervention in primary care and experiences of dementia care during COVID-19. | Topic guide explored: key changes to dementia care during COVID-19, including changes in caseload and role; impacts of COVID-19 on staff and service users. | Topic guide explored: views of experts on the annual dementia review; factors influencing successful review; and examples of good practice. |
| Date of data collection | March-August 2019 | July-December 2019 | November-December 2020 | February-May 2021 | August-November 2019 |
| Data management and confidentiality | Interviews and focus groups were audio-recorded, transcribed and checked and pseudonymised for analysis. Most participants provided written consent via email or post, with verbal consent sought before interview for those who had not returned consent forms. Participants are identified in this paper via pseudonym. |  |  |  |  |
| Ethical approval | NHS Research Ethics Committee Wales 3 (reference 18/WA/0349) |  |  |  | Newcastle University Faculty of Medical Sciences Research Ethics Committee |
| Authors responsible for data collection & analysis | CB, GB, AW | CB, GB, AW | CB, GB, AW | AW, MP | JD, AW |

### Integrated dataset

Relevant data from PriDem and ADR were integrated and analysed thematically [27] (CB, GB, AW); a pragmatic, ‘codebook thematic analysis’ approach was chosen for its utility in efficiently exploring specific information needs (i.e. data relating directly to annual dementia review) within such a large dataset [28]. Themes from data from service users and professionals were derived separately before being combined, to better capture similarities and differences.

## Results

### Participants

Participants are shown in Table 2. A total of 209 individuals took part (some participated in more than one part of the dataset), including 48 service users. Of the professional participants, 41 were based within primary care, including 18 GPs; others included a range of professionals from secondary care, social care services and the third sector across England and Wales.

**Table 2:** Overview of participants

| Participants | WS2.<br>1 | WS2.<br>2 | WS3.<br>1 | WS3.<br>2 | ADR | Unique<br>participants |
| --- | --- | --- | --- | --- | --- | --- |
| People living with dementia | 0 | 17 | 3 | 0 | 0 | 17 |
| Carers | 0 | 31 | 10 | 0 | 0 | 31 |
| Commissioners | 25 | 0 | 4 | 6 | 0 | 26 |
| Service managers | 25 | 9 | 8 | 8 | 0 | 31 |
| Frontline staff | 11 | 75 | 13 | 7 | 0 | 91 |
| Experts | 0 | 0 | 1 | 0 | 13 | 13 |
| **Total** | **61** | **132** | **39** | **21** | **13** | **209** |

### Views on annual dementia review

Themes developed from the data were:

- Perceived benefits of annual dementia review
- Variability and (in)visibility of annual dementia review
- Logistics of annual dementia review
- External influences and constraints on annual dementia review

Both professionals and service users highlighted potential benefits of annual dementia review but also raised concerns about how such reviews were performed in practice. There were differences in nuance between the two participant groups; points of comparison and contrast are described in more detail in the following sections.

#### Perceived benefits of annual dementia review

For service users, key potential benefits included the opportunity to engage in a proactive, holistic conversation exploring their needs, and receive support and reassurance in understanding symptoms and changes. This proactive element was especially important as service users were not necessarily aware of the range of symptoms that could be related to dementia, and many reported difficulties in accessing GP appointments or were otherwise reluctant to contact their GP:

> *good idea, that [annual dementia review] […] I don’t talk to people about my troubles and things like that. If a doctor was asking you how you are and all the rest of it, then you would open up to him, but I think you need somebody to talk to. If somebody comes around and asks you these questions, (Laughter) one thing leads to another. (P604, person living with dementia)*
>
> *It might be presenting as dementia when they keep getting up or something like that, but it might be a pain that’s affecting them that’s nothing to do with dementia. So that [review] would be a very good thing. (C301, carer)*

Both frontline professionals and commissioners similarly identified annual dementia reviews as an important ‘safety net’ for service users who were not actively seeking help which could allow problems to be identified at an early stage:

> *[…] where they work well, people value them very highly and they do tend to pick up issues and problems before they become more severe so they almost certainly are very helpful in reducing stress, anxiety and other problems for the person with dementia and their carers and probably do help to reduce unnecessary hospital admissions (ADR S11, dementia specialist nurse)*

Not all participants, however, were convinced of the value of an annual dementia review in primary care. Some experts suggested that an annual dementia review alone was not sufficient, and that it should be part of a broader overall intervention such as case management:

> *[…] annual review standing alone doesn’t necessarily meet all the needs of the patient. That’s the problem. So they need further access to care in between annual reviews (ADR S3, geriatrician)*

Service users questioned whether GPs would have the skills and knowledge to conduct dementia reviews and the interest and willingness to take on this care:

> *Would he [GP] be qualified enough to do that? [Interviewer: What do you think?] No. He’ll know a lot, but he probably won’t know what the specialist knows. […] I don’t know of any GPs who are up to that standard with Alzheimer’s and dementia (P004, person living with dementia) I can’t see what he [GP] can do. So more often, what can you do? You can’t do anything, can you? […] it doesn’t seem as if they’re interested or if they can help. (P005, person living with dementia)*

This often reflected a broader lack of awareness by both service users and professionals of what kinds of support were available and a lack of confidence that anything could be done to help people living with dementia. Participants emphasised the importance of creating a clear action plan following review, in order for service users to feel that the review had tangible benefits and that support was available:

> *[…] when we were just seeing the GP, you see, I just felt like we were reviewed, limbo land, reviewed, limbo land. Nothing happened. […] Whereas, when [specialist dementia clinician] came, I thought that we moved on. We had things done. We had a CT [scan]. She put medication in place. We started to be proactive in his care and though we know that there’s not going to be any cure, at least it felt like somebody was taking hold of what was happening and got somebody who was going to act on his concerns. (C404, carer)*

#### Variability and (in)visibility of annual dementia review

While several service users recalled having had reviews for other conditions, for example a wellness check or diabetes review, most service users were unaware of having had a specific annual dementia review:

> *I can honestly say we’ve never had a dementia [review]… Whether it’s done via the diabetic follow-up, I don’t know. He obviously has a diabetic follow-up every six months […] We’ve never had, “This is your annual review from dementia*.*” (C202, carer)*

Moreover, where dementia symptoms had been explored during a generic review, some carers perceived this as lacking depth. Professionals similarly raised concerns over the extent to which annual dementia reviews were taking place. One potential reason for perceived low rates of completion related to the lack of skills, knowledge and confidence of GPs in dementia, particularly in prescribing dementia medication:

> *[…] our previous GP chair for [this area] said that he couldn’t understand why GPs weren’t willing to do [annual dementia reviews] […] they do reviews for lots of other physiological medications that they don’t normally work with, that they’re not familiar with. So he couldn’t understand why people [GPs] were singling out dementia. (S030, NHS commissioner)*

Another factor related to the accuracy of practice dementia registers; inconsistencies with the coding of dementia diagnoses could lead to some patients not receiving reviews or follow-up:

> *[Some] patients have a diagnosis of dementia, yet they’ve not all been coded correctly. […] you can miss out on patients and then patients can also miss out on support and care, if you’ve not got them properly within EMIS [primary care computing system]. (S306, GP)*

While most participants felt that reviews should be holistic, considerable variability in the quality of reviews was reported: in some areas, ‘enhanced’ dementia reviews were commissioned which were more detailed and linked with other local dementia services; in other areas, reviews were perceived as ‘cursory’ by professionals in other services. Key elements flagged by professionals as frequently missing from annual dementia reviews included swallowing, carer health, and planning and goal setting. Other elements important to service users included medication review and support for carers, although the latter could be complicated if carers were registered with a different GP practice.

A barrier to holistic dementia reviews was time, although there were elements (e.g., physical health checks) that could be task-shared with other professionals or combined with other reviews. Some professionals argued that multiple reviews for LTCs should be replaced by a single holistic review of which dementia was one element; one successful example of this was in a GP practice where reviews were conducted by a specialist with experience of both dementia and physical health:

> *[We’re] trying to have a, sort of, one review for everything. So, rather than drag people in for a diabetes review, drag them in for a COPD review, hypertension review, dementia review, them having to come in and see loads of different people, we’re trying to train people up […] so that [they] can undertake all those other reviews when [they go] to see someone to try and prevent duplication. (S403, GP)*

#### Logistics of annual dementia review

Discussion of logistics of annual dementia reviews covered timing and frequency, location, and responsibility for review. Opinions on the timing of the first review ranged from two weeks to six months after diagnosis, and on frequency of subsequent reviews included every three, four, six or 12 months. In one area, additional reviews by GPs had been commissioned in order to increase parity with other conditions:

> *[…] within the QOF, GPs have an annual review of everybody with dementia, and their carers. We commissioned another, because we thought a year is a bit long. And if you compare it with any other long-term conditions, you have [reviews] much more often (S021, NHS commissioner)*

However, in areas where other dementia support was in place, for example a local telephone helpline, dementia navigator, or follow up from memory clinic, less frequent reviews by GPs were acceptable to service users:

> *I think once a year is alright for now, for me, once a year. I’ve got [access to a dementia navigator] in between. […] So I’m sort of covered, really, for the whole year. (P203, person living with dementia)*

The lack of consensus around frequency suggests that there is a strong need for flexibility in timing of reviews, tailoring to stage of dementia, speed of progression, other support, and service user preference.

While the value of face-to-face contact was emphasised, GP practices could be difficult to access, particularly for people with more advanced dementia, making home visits necessary. The importance of continuity was also highlighted; while some service users reported positive, ongoing relationships with their GP, others reported that they did not see the same GP at each visit and therefore felt they were not the professional who knew them best:

> *Well, the trouble is there are about six, seven or eight of them [GPs], and you see a different one each time you go, and I don’t go very often anyway. And so you never know really who you’re talking to. (P201, person living with dementia)*

A range of professionals, either within or external to GP practices, was reported to be involved in contributing to or directly providing annual dementia reviews. An example of the latter was the commissioning of GPs with an extended role (GPWERs) to provide enhanced reviews of all people living with dementia across a local area. By providing reports and flagging actions for the patient’s usual GP, the GPWERs contributed information which could be used in completing an annual dementia review. This was valued by local GPs:

> *[GPWERs send us] actions, as well, if, you know, ‘I’ve started the patient on this medication, please can you prescribe it,’ or bits and pieces like that. If there’s something extremely urgent, as well, they’re really good at contacting us via the phone […] if you need to have access to their [GPWERs] expertise you can do that (S306, GP)*

Similar ongoing follow-up and review was also undertaken by memory services in some areas. Where such services were in place, GPs reported that this helped to cover people who had not been seen in general practice. However, where systems were not well joined up, there could be duplication and weak co-ordination between reviews. There were also examples of GP practices where reviews were largely the responsibility of a co-located dementia specialist clinician (frequently a nurse or allied health professional). Potential advantages of this approach included improved continuity, dedicated time for conducting dementia reviews, and reduced pressure on GPs. Service users valued the flexibility this approach offered:

> *I want to talk about things. […] They [GPs] have a [limited time] slot, but with [specialist dementia clinician], you don’t get that feeling. Maybe he’s just learnt how to be*… *He’s a very friendly person, and understanding. (P006, person living with dementia)*

Rather than appointing a specialist clinician, some participants suggested that a cost-effective solution was for existing members of the primary care team, such as advanced practitioner nurses, to take on dementia reviews.

#### External influences and constraints on annual dementia review

Factors influencing the conduct of annual dementia reviews included the practice context and setting; the COVID-19 pandemic; and the structure and constraints of the QOF itself. Some professionals attributed variability in quality and consistency of reviews to capacity within practices, with a key factor being time, leading to lower priority being given to dementia reviews:

> *I think it depends on the individual doctor, the individual surgery, where you’re located in the country, the social class of the area. I live in a really nice area so the doctors have loads of time for us, there’s hardly anybody there, they come out and greet you by name. But I work in the middle of [a city] where the doctors are overloaded […] I’m sure that’s why when you might go for an appointment about your foot it ends up being your dementia review and that’s ticked the box (ADR S9, chief executive, third sector care provider)*

The COVID-19 pandemic exacerbated existing problems with annual dementia reviews. Commissioning and primary care resources had frequently been reassigned to deal with COVID care and vaccination, leading to pauses or reduced focus on service development and quality improvement, including annual dementia review. Scheduled annual dementia reviews were not consistently taking place during COVID, raising concerns about potential adverse outcomes for service users and negative consequences for other services. Some GPs tried to address the lack of planned reviews during COVID by conducting opportunistic dementia reviews for those who attended for other reasons:

> *[…] we haven’t been deliberately bringing people in just to do routine things. We’ve tried to make the most of when we do have to see them, we try and do the routine stuff as well, so you make the most of every face-to-face contact you have. So we wouldn’t bring somebody in to do an annual dementia review, but if we were seeing them because we needed to see them, we would try and make sure we covered those areas as well. (S043, GP)*

While financial incentives from the QOF were identified as a motivating factor for ensuring reviews take place, several participants raised concerns that this contributed to reviews being seen as a ‘box ticking exercise’ rather than a thorough review in which service users were actively involved. The usefulness of the QOF indicator for dementia was queried by participants, who argued that dividing the QOF points over a greater number of aspects of ongoing care and management, as is the case for some other LTCs such as diabetes, would improve review quality:

> *[…] it’s a tick box exercise. I don’t think tick box exercises are necessarily that bad if you compare it to doing nothing at all which is what happened before […] [but] are there things that we could have the 39 points rather than just saying, ‘You get one for’ – sorry, I think the 39 is for the whole of dementia. […] It’s a bit of a blunt instrument. (ADR S12, old age psychiatrist)*

Moreover, although the potential for providing holistic support was thought to be a strength of conducting dementia reviews within primary care, there were concerns that the QOF structure encouraged non-holistic working by reimbursing different LTCs separately. Several experts felt that a standard approach to reviews for other LTCs had been adopted without consideration of how this needed to be adapted for dementia:

> *[…] dementia is more interesting than diabetes, so by definition it’s a bit more complicated. I mean for diabetes presumably, it’s look at the blood sugar, see if they’ve got a neuropathy and make sure that the kidneys and eyes are alright so it’s relatively simple, because it’s formulaic you could do it very simply. Dementia is a bit more nuanced […] we don’t have a blood test that we can measure or anything as simple as that. (ADR S12, old age psychiatrist)*

Both service users and professionals highlighted the need for flexibility in tailoring the content of the review according to the needs of the individual, which the use of strict templates could make problematic if the professional carrying out the review did not have confidence in providing dementia care:

> *[…] it’s become much more of kind of a medical tick box exercise rather than a quality intervention if you like, so they’re asking the questions but it’s like it’s coming off a script. […] it relies on the person who’s doing the review to have an understanding of what else to ask for and what they’re actually looking for and my experience is a lot of the practice staff don’t know what the red flags would be to actually do anything about the situation that the person with dementia or the family carer is experiencing. (ADR S6, dementia specialist nurse)*

## Discussion and conclusions

Despite high GP participation in the QOF in England and Wales, and high completion rates for the dementia indicators including annual dementia reviews, this study raises critical questions about the quality and content of such reviews, even before the COVID pandemic. Our data suggest that people living with dementia and carers may not be aware of an annual dementia review taking place, with some professionals expressing similar concerns over completion, process and quality of reviews. Both service users and providers considered the annual dementia review potentially beneficial, providing an opportunity for proactive, holistic and personalised care and acting as a safety net for those who may not otherwise attend; however, some professionals questioned whether GPs had the capabilities and/or capacity to undertake reviews effectively. The structure, and constraints, of the QOF itself were seen as influential factors; professionals suggested financial reimbursement should be based on both quantity and quality, including a range of aspects of care and management, as is the case for other LTCs, with the content of reviews more effectively specified. However, such indicator would also need to be sensitive to the importance of tailoring to individual needs; for example, a review which focused on one issue in-depth may be of high value to a patient but could be difficult to capture with an indicator.

Our findings are consistent with earlier studies which questioned the quality and comprehensiveness of annual dementia reviews [17, 18]. Between 2006 and 2013, a UK-wide study reported that only half of people living with dementia had a documented annual dementia review [17]; between 2008 and 2009, a smaller study of GP practices in Manchester found that 80% of people living with dementia had a documented annual dementia review [18]. Both studies commented on the variable quality of reviews with limitations around integration of other LTCs, documentation of discussions with carers, social care review, and inappropriate prescribing of antipsychotics. Previous evaluations of QOF in general suggest that ‘tick box’ exercises could lead to poorer patient experiences [8] and that QOF as a ‘pay for performance’ scheme led to dilemmas among primary care professionals as to whether to focus on income and targets, as opposed to patient-centred practice [29].

While QOF mechanisms were paused during COVID-19 lockdowns, some remote ‘check up’ calls were reported by GP practices; other research has highlighted problems with such remote consultations including reduced quality and access difficulties [30, 31].

Positive strategies for enhancing annual dementia reviews including involving other members of the practice team (e.g. nurse, pharmacist), working collaboratively with other services to share information, and integrating dementia reviews with other LTCs to ensure an efficient, holistic approach. In order to better support families affected by dementia, it is important to include clear information and signposting that a dementia review is taking place, and to ensure that they are given space to talk about matters that concern them. Timing, frequency and location of reviews should be flexible to best accommodate individual needs and preferences [11]. Service users emphasised the importance of continuity, echoing findings from recent research which found GP continuity in dementia care was associated with safer prescribing and lower rates of major adverse events [32]. Concerns about GP capacity and capabilities to undertake reviews resonate with recent evidence about task-shifting post-diagnostic support to primary care [13] and the search for more effective service models [33]. While capacity and capabilities are distinct issues, some solutions, such as embedding clinical dementia expertise within primary care, can contribute to addressing both: capacity in the short term and capabilities in the longer term through opportunities for learning and awareness raising [13].

### Strengths and limitations

A strength of this study is the varied and large number of participants, ensuring that a broad range of perspectives was incorporated, including those of GPs with experience of providing annual dementia reviews and, importantly, people living with dementia and their families.

Data were collected between 2018 and 2021, enabling changes due to the COVID-19 pandemic to be captured. A potential limitation is that the PriDem work was not specifically focused on annual dementia reviews, with the result that data may be less detailed than a study purely investigating this topic. Most service users had little personal experience of an annual dementia review which limited their ability to comment; however, this does demonstrate problems with the current system where service users are often not clear that a review has taken place. Many professionals interviewed worked outside primary care and therefore often reported perceptions of annual dementia review rather than direct experience. Nevertheless, the data contain useful insights and the parallel themes identified by service users and professionals suggest they are robust.

### Recommendations for practice and future research

Despite the annual dementia review being part of the QOF framework since 2006, there is limited data about its administration, acceptability or effectiveness. Our findings echo those of others suggesting an urgent need for the development of more nuanced and detailed QOF dementia indicators to provide formal structure and improve quality and service user satisfaction. Involving service users and frontline practice teams in developing such indicators may help ownership and participation [34], thereby ensuring that the review is person-centred and deliverable. Learning lessons from the management of other LTCs may be helpful [35]. A combination of training and collaborative work, for example through training a lead GP or practice nurse in dementia care or task-sharing with professionals with specialist expertise [36], could help to address concerns about the both the capacity and capability of primary care to undertake such reviews and may provide a framework for the task-shared approach urgently recommended for implementation on a wider, global scale [4]. We recommend future research explores experiences of annual dementia review for under-served populations (e.g. people with young onset dementia and with rare dementia subtypes; people from minority ethnic groups) and examines templates currently used for dementia reviews and resulting care plans as part of developing a standardised model for a more detailed, holistic, and effective annual dementia review.

## Data Availability

PriDem data are available upon reasonable request via

https://doi.org/10.25405/data.ncl.c.5718116

## Acknowledgements

Administrative support was provided by Angela Mattison. We thank all participants for their support and involvement in the study. The PriDem study team is led by Professor Dame Louise Robinson (Newcastle University) and also includes Louise Allan (University of Exeter), Sube Banerjee (University of Plymouth), Alistair Burns (NHS England and NHS Improvement), Sophie Dimitriadis (International Longevity Centre), Karen Harrison Dening (Dementia UK), Sarah Griffiths (University College London), Derek King (London School of Economics), Martin Knapp (London School of Economics), Doug Lewins (PPI co-applicant), Jill Manthorpe (King’s College London), Greta Rait (University College London), Emily Spencer (University College London), Sue Tucker (PPI co-applicant), Kate Walters (University College London), Jane Wilcock (University College London) and Raphael Wittenberg (London School of Economics).

## Data availability statement

PriDem data are available upon reasonable request from data.ncl: https://doi.org/10.25405/data.ncl.c.5718116

